# Neonatal admission as a marker of risk for poor educational attainment and special educational needs in children aged 5-11 years

**DOI:** 10.64898/2026.07.15.26358132

**Authors:** Abinayah John, Cleo Pike, Laurentya Olga, Ulla Sovio, Hilary S Wong, Gordon CS Smith, Catherine EM Aiken

## Abstract

**Background:** Children born prematurely (before 37 weeks) or admitted to the neonatal unit (NNU) are at increased risk of adverse long-term physical health outcomes. It is also recognised that there is an association with later academic performance and special educational needs, however it is not clear whether these broad risk factors could be used as stand-alone heuristics to identify children who may benefit from additional support in educational settings.

We aimed to examine the associations between neonatal unit (NNU) admission and educational attainment in mid-childhood.

**Methods and Findings:** Pregnancy data from a prospective birth cohort (Pregnancy Outcome Prediction Study, Cambridge, United Kingdom, 2008–2012) were linked to national educational outcomes (Department for Education, United Kingdom). Multivariable regression models adjusted for maternal, child, and socioeconomic factors were used to evaluate associations between (i) all NNU admissions, (ii) at term NNU admissions >48 hours, (iii) preterm birth without ongoing physical health needs, and educational outcomes at ages 5–11 years.

Children who required any NNU care were more likely not to meet expected educational standards across multiple ages and domains in early and mid-childhood: age 5 early year foundation (aOR 1.64, 95% CI 1.19–2.27, p=0.003), phonics at age 6 (aOR 2.43, 95% CI 1.72–3.57, p<0.001), and at age 7 (here assessments were divided into multiple domains): reading (aOR 1.67, 95% CI 1.18–2.38, p=0.004), writing (aOR 1.72, 95% CI 1.25–2.38, p<0.001), mathematics (aOR 1.56, 95% CI 1.09–2.22, p=0.020), and science (aOR 1.85, 95% CI 1.22–2.78, p=0.003). Similar patterns were observed among both at term-born infants who stayed >48hrs in NNU (phonics assessment at age 6 aOR 2.26, 95% CI 1.51–3.36, p<0.001) and in children born preterm without long-term physical health sequelae (phonics assessment at age 6 aOR 3.07, 95% CI 1.96–4.81, p<0.001). These associations were robust to adjustment for demographic, perinatal, and socio-economic factors. By age 11, differences in academic attainment were attenuated and no longer clearly distinguishable across all exposure groups. However, there was an increased likelihood of special educational needs (SEN) at age 11 associated with any NNU admission (aOR 1.78, 95% CI 1.15–2.73, p=0.009), at term NNU admission for >48hrs (aOR 1.88, 95% CI 1.19–3.00, p=0.007), and children born preterm without long-term physical health sequelae (aOR 1.50, 95% CI 1.00–2.25, p=0.049). Predictive performance of any NNU admission for SEN at age 11 was moderate (AUC 0.70, 95% CI: 1.14-2.65, p=0.010), with balanced sensitivity and specificity and high negative predictive value.

**Conclusions:** NNU admission, for both term and preterm infants, is associated with poorer educational outcomes and an increased likelihood of special educational needs in mid-childhood.

## INTRODUCTION

In recent years there has been growing interest in the long-term consequences of early-life medical complexity, including the effect of those complexities on later health and developmental outcomes. Low birthweight and preterm births are associated with lower attainment on educational benchmarks across global contexts [1–4], and children affected are more likely to achieve lower scores on cognitive tests [4–8]. Neonatal unit (NNU) admissions are rising worldwide as the heterogeneity of maternity populations and the efficacy of interventions offered increases [9].

Approximately 1 in 10 babies in the UK requires NNU admission, with ∼70,000 infants admitted each year [10, 11]. However, while follow-up frameworks such as NICE guidelines target specific high-risk groups (e.g. very preterm infants), the majority of neonatal admissions fall outside these pathways and are not followed up within a comprehensive national framework [12]. The magnitude of educational risk in this broader population remains unclear. Moreover, given resource constraints, these children are not systematically followed into school age.

The ability to link early life medical factors to educational outcomes is a crucial aspect of understanding which children are likely not to meet nationally-benchmarked educational standards. The UK data environment, including the Secure Research Service provided by the Office of National Statistics, and the collection of standardised educational outcomes makes this a tractable aim. Our previous work demonstrates feasibility for these linkages within research cohorts [13] using detailed medical information. Here we took the approach of determining whether health-based heuristics that are readily ascertainable by parental report and through medical health records could identify a child as at risk for poor educational outcome. We aimed to determine whether any NNU admission alone could predict educational outcomes during mid-childhood (ages 5-11 years), or whether it may be a good candidate risk factor in combination with other metrics to attract additional early learning support.

## METHODS

### Study population and data sources

The Pregnancy Outcome Prediction Study (POPS) is a prospective UK birth cohort that recruited unselected nulliparous women with singleton pregnancies from the Rosie Hospital, Cambridge, United Kingdom, between January 2008 and July 2012 (N=4212). The full study design has been reported in detail elsewhere [13–15]. Recruitment took place typically before 14 weeks of gestation following confirmation of a single viable fetus via ultrasound scan. Crown-rump length measurement was used to determine the dating of the pregnancy. Antenatal delivery, and neonatal outcome data were collected by the research team from questionnaires completed by study participants.

Additional data were extracted from paper and electronic medical record systems. We linked antenatal and neonatal phenotypic data to healthcare utilisation (Hospital Episode Statistics data via NHS England) and educational outcomes (National Pupil Database via the UK Department for Education). For the current analysis, POPS participants who had live-born singleton infants were eligible for inclusion. There were 3722 mother/baby pairs who were confirmed alive and traceable at study commencement. 45 pairs opted out; therefore, linkage was attempted for 3677 (88% of eligible cohort; Fig 1).

**Fig 1:**
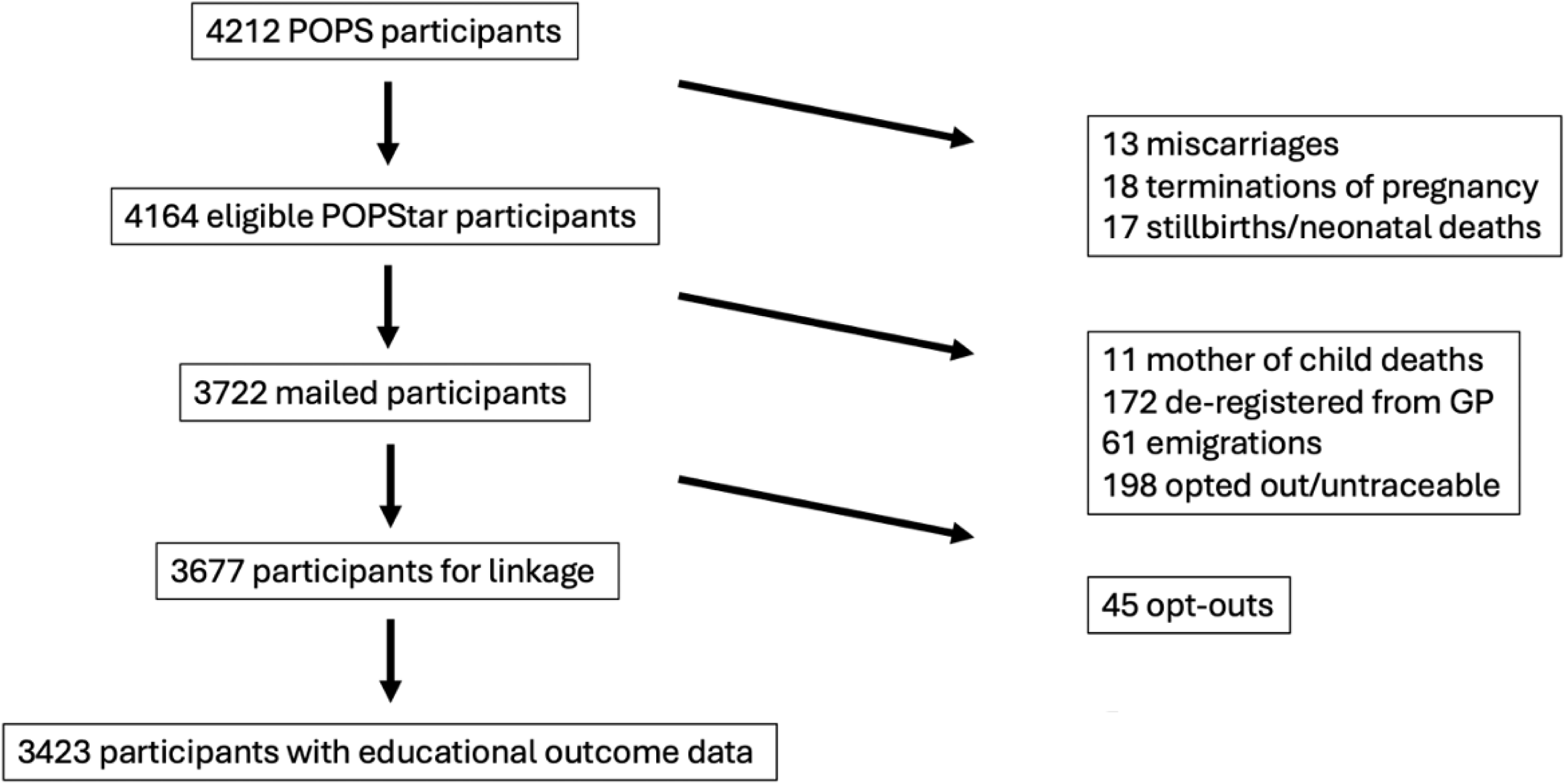
Number of participants from recruitment to Pregnancy Outcome Prediction (POP) study through identification of the analytical sample. Flow diagram shows participant eligibility, linkage availability, and inclusion in the analytic cohort. The analytic sample comprised all children with POPS cohort data and at least one linked educational outcome at any assessment time points between ages 5 and 11 years (N=3423). Children without linked data at age 5 could still contribute data at later assessment points if successfully linked; therefore, the analytic cohort was defined using availability of any educational outcome rather than complete data at all time points.

We defined a referent group of children as those who were: (i) born with birthweight between the 10th and 90th centiles (all birthweight centiles were based on the relevant 1990 UK population growth reference [16] adjusted for gestational age and sex and categorised into predefined centile groups), (ii) born between 37+0 and 41+0 weeks’ gestation, and (iii) did not require NNU care. This referent group was compared with predefined analytic groups comprising children with (i) any NNU admission, (ii) NNU admission for >48 hours at term, and (iii) preterm birth (<37+0 weeks’ gestation) without ongoing physical health sequelae, defined as the absence of pre-specified childhood morbidities ascertained via Hospital Episode Statistics data (S3 Text). A threshold of >48 hours was applied to term NNU admissions to distinguish clinically significant neonatal illness requiring sustained intervention from brief admissions for observation or transitional care. This threshold was pre-specified in the statistical analysis plan (see S1 Text). Restriction to term-born infants additionally allows the contribution of neonatal illness to educational outcomes to be examined independently of prematurity, which is itself strongly associated with later developmental outcomes.

### Exposures and covariates

The primary exposures were any NICU admission, NICU admission for >48 hours at term, and preterm birth.

Covariates were selected based on biological plausibility and data-agnostic approach. Covariates included in the analysis were maternal age (in years), pre-pregnancy maternal BMI (derived from maternal weight at booking divided by the square of measured height (in kg/m^2^), maternal ethnicity (White/European vs non-White), smoking history during pregnancy (yes/no), partner status (yes/no), maternal occupation (4 categories: (i) managerial or professional occupations, (ii) associate, administrative and skilled occupations, (iii) unskilled occupations, (iv) freelance/self-employment excluding business owners)/unemployed, infant sex (male/female), season of birth (born in winter, spring, summer, or autumn), index of multiple deprivation (IMD score; 2007), school funding type

(academy, community, or voluntary; standard models of UK state school funding), school year at assessment, and presence of serious childhood morbidity. Significant childhood morbidity was defined as any clinically relevant childhood illness or medical condition not considered to arise from intrauterine developmental processes and therefore treated as a potential confounder in analyses of perinatal factors and mid-childhood educational attainment (S1 Table; [12]). Gestational age at birth was additionally included as a covariate in models examining any NNU admission and NNU admission >48 hours at term, but was excluded from models examining preterm birth as the exposure, since gestational age was used to define this exposure group. These data were extracted from the Hospital Episode Statistics (HES; provided via NHS Digital) relating to the emergency department attendances, hospital inpatient stays, and hospital outpatient appointments. The definitions of the individual covariates are reported in further detail in S1 Text.

Educational outcomes were derived from the National Pupil Database; standardised national assessments conducted within the UK school curriculum. At age 5, school readiness was assessed using the Early Years Foundation Stage profile, a national measure of early childhood development and preparedness for formal education. At age 6, children completed the national phonics screening check, which evaluates early reading and decoding ability. At age 7, corresponding to primary school year 2 (Key Stage 1), attainment was assessed across reading, writing, mathematics, and science. At age 11, corresponding to the end of primary school (Key Stage 2), outcomes included reading, writing, and mathematics. For all assessments, outcomes were defined relative to nationally benchmarked expected standards, allowing comparison with the wider population of children attending UK state schools (see S2 Text).

At age 11, educational outcome data were available for a smaller subset of the cohort compared with earlier time points because national Key Stage assessments in England were disrupted during the COVID-19 pandemic. In particular, Key Stage 1 and Key Stage 2 assessments were cancelled in 2020 and 2021, resulting in missing attainment data and reduced linkage availability for some children within the cohort. Consequently, analyses at age 11 were based on smaller sample sizes, leading to wider confidence intervals and reduced statistical power.

Special educational needs (SEN) status was derived from the National Pupil Database and reflects provision recorded within the school system, including SEN support and Education, Health and Care plans. SEN data were only available at age 11 within the linked dataset and were therefore not analysed at earlier time points.

### Statistical analyses

The analysis followed a prespecified statistical analysis plan (see S1 Text). Baseline demographics were described using numerical and categorical variables, presented as mean (±SD) and number (percentages), respectively. The associations between exposure groups and educational outcomes were modelled using multivariable logistic regression, adjusted for identified covariates. Covariates that had a small proportion of missing values, including maternal BMI, partner status, and school funding status were imputed using multiple imputation by chained equations (MICE) under a “missing-at-random” assumption [17]. The R package “mice” (3.16.0) was used to generate 20 imputed data sets, using linear regression for continuous variables and logistic regression for categorical variables. The following variables were included in the imputation models: GA, birthweight centiles, delivery modes, maternal factors (age, BMI, occupation, ethnicity, partner status, alcohol consumption, smoking status), IMD, school funding type, and educational outcome z-scores at the age of 5 years (detailed elsewhere/included in S2 Text). The gestational (GA) was used in imputation models for all exposure groups but was only included as an adjustment covariate in the regression models for any NNU admission and NNU >48 hours at term, not in models examining preterm birth as the exposure. Analyses run on each data set were pooled according to Rubin’s rules [18], and imputed values were found to be comparable to observed values.

Sensitivity analyses were pre-specified to assess the robustness of findings. These included: (i) evaluation of model discrimination using receiver operating characteristic (ROC) curves and area under the curve (AUC); (ii) stratified analyses by child sex, examining both univariate and multivariable associations between controls and each defined risk group; (iii) repeat analyses excluding preterm births from risk groups not directly related to prematurity; and (iv) exclusion of children with severe childhood morbidities, defined as previously described (Olga et al., 2023).

All analyses were conducted using R 4.4.0 (R Core Team, Vienna, Austria) [19]. Two-sided P values are reported. Reporting of this study is compliant with the STROBE (Strengthening the Reporting of Observational studies in Epidemiology) guidelines.

Written consent was obtained from all participants prior to the antenatal study. Participants were informed about planned data linkage and retained the right to withdraw at any point. Individuals were excluded automatically if NHS England did not hold valid updated contact information. Data linkage was performed in anonymised form, with access provided through the Office for National Statistics Secure Research Service, where datasets were analysed using randomly allocated study identifiers only. Written ethical approval was granted by the Cambridgeshire 2 Research Ethics Committee (07/H0308/163), the Cambridge Central Research Ethics Committee (18/EE/0036), and the Confidentiality Advisory Board (18/CAG/0024).

## RESULTS

The analytical sample included ∼81% (3423/4212) of the original birth cohort (Fig 1). The baseline demographics of the analytical sample were comparable to those of the entire POPS cohort described elsewhere [13–15] (S3 Table).

### Associations between NNU admission and mid-childhood educational attainment

There was a significant difference in the likelihood of not meeting expected educational standards at ages 5, 6, and 7 between children who had experienced any NNU care and those who had not (Fig 2, S2A Table). This effect persisted throughout early and mid-childhood (ages 5–7) and was consistent across all domains assessed (reading, writing, science, and mathematics).

**Fig 2:**
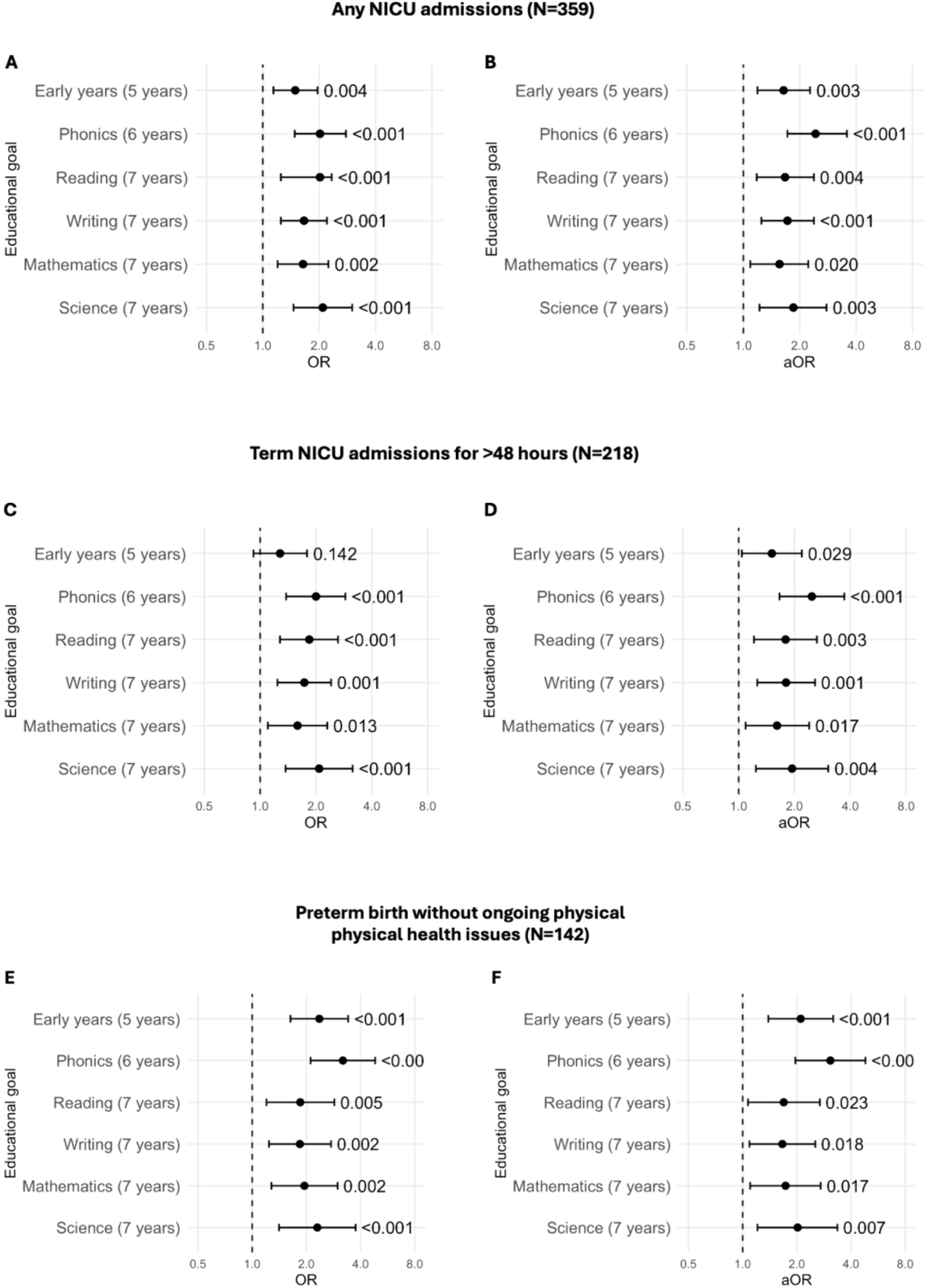
Association between NNU admissions and childhood educational attainment at age 5 to 7. All panels: OR with 95% confidence intervals (Cls, plotted on a log2 scale) of educational attainment, with the referent group defined as children not admitted to NNU, born >10th and <90th centile, and born at term. Panels A,C,E: unadjusted results. Panels B, D: adjusted for maternal factors (age at pregnancy, ethnicity, occupation, partner status, smoking history), infant factors (GA, sex, birthweight standard deviation score, birth seasonality, childhood physical health), and socioeconomic factors (index of multiple deprivation, school funding type, academic year). Panel F: adjusted for the same covariates excluding GA, since GA was used to define the preterm exposure group. Panels A&B: any NNU admission (N=331). Panels C&D: term NNU admission for >48 hours (N=218). Panels E&F: preterm births without ongoing physical health issues (N=142). The complete-case sample varies across panels and educational stages, as not all children in the analytic sample had linked outcome data available at every assessment timepoint. OR, odds ratio; aOR, adjusted odds ratio; NNU, neonatal unit; GA, gestational age.

Similar patterns were observed when analyses were stratified by pre-specified NNU sub-groups (Fig 2, S2A Table). Children born at term who required NNU care for more than 48 hours had increased odds of not meeting expected standards at ages 5–7, with effect estimates comparable to those observed in the overall NNU cohort. Children born preterm and without ongoing physical health needs during childhood also demonstrated increased likelihood of poorer attainment across early educational domains, with effect estimates generally larger than those observed for NNU admission alone.

At age 7, the association between any NNU admission and failing to meet educational standards remained evident across reading, writing, mathematics, and science. For children born at term and with neonatal unit admission for >48 hours, we observed increased odds were observed across all domains, although the association for mathematics was attenuated following adjustment. For children born preterm and with no ongoing physical health needs, we observed increased odds across all domains, both before and after adjustment.

Further analysis demonstrated that there were no significant differences in the impact of NNU care between male and female children (S2B Table). In addition, findings were persistent across ethnic groups and socio-economic strata (as proxied by index of multiple deprivation), and persisted after adjustment for school characteristics including funding type and academic year (Fig 2).

### Associations of NNU admission and preterm birth with educational attainment at age 11

Differences in expected academic attainment at age 11 were smaller and less consistent across all exposure groups (Table 1). For any NNU admission and term NNU for >48 hours, point estimates remained generally above 1, but adjusted associations were attenuated and no longer statistically significant for reading and mathematics; the association with writing remained of borderline significance for any NNU admission only. For children born preterm without ongoing physical health issues, adjusted associations remained statistically significant for both writing (aOR 1.92, p=0.042) and mathematics (aOR 2.11, p=0.018), indicating a more persistent effect of prematurity on attainment at age 11 compared with NNU admission alone.

**Table 1:**
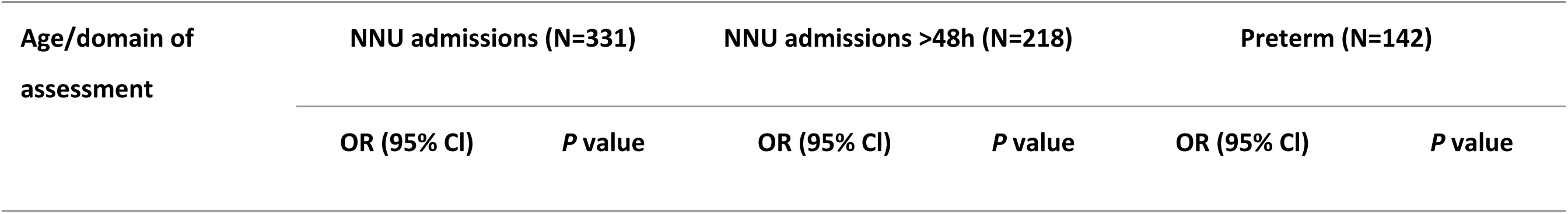
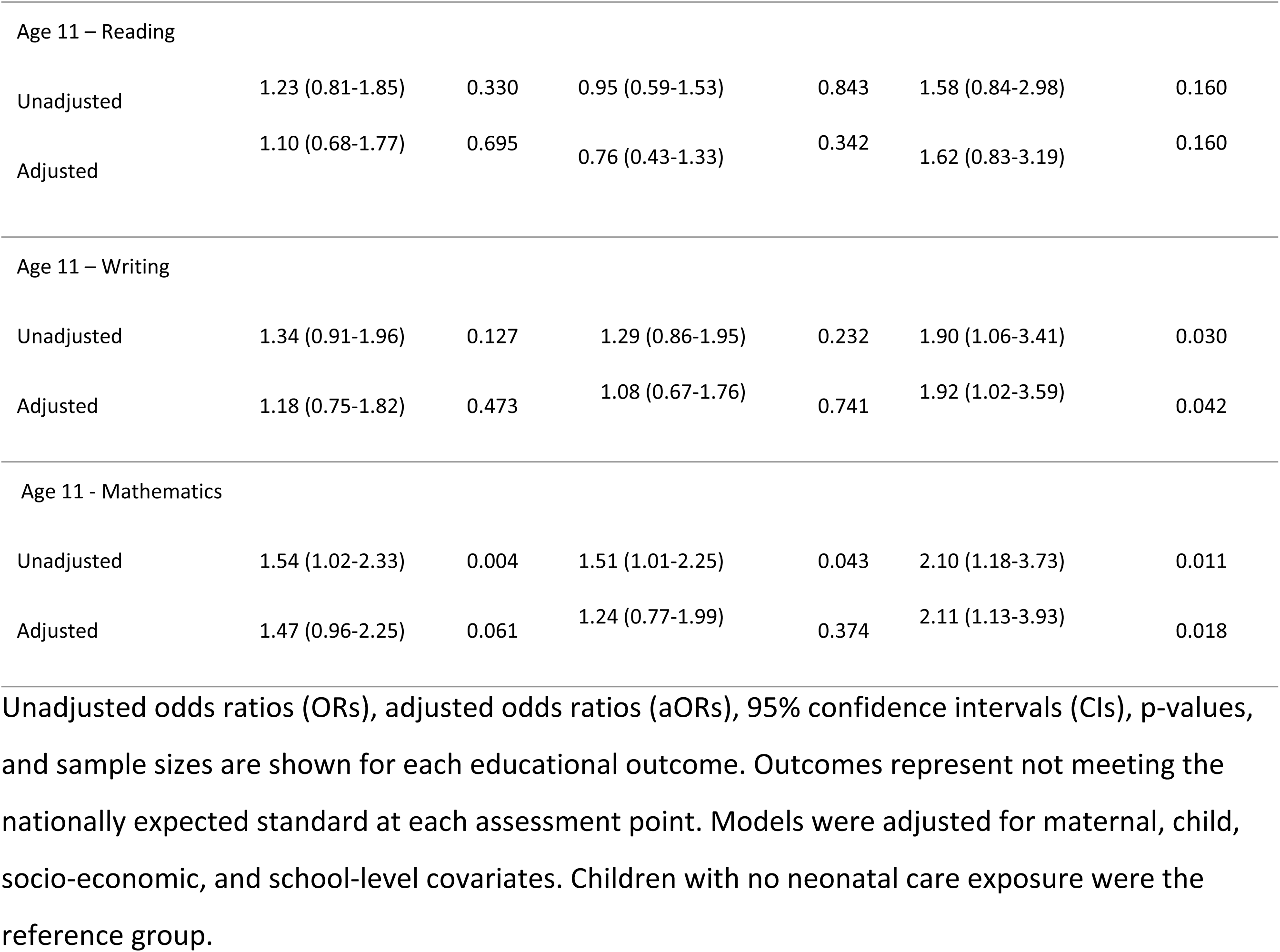
Association between NNU admissions and childhood educational attainment at age 11.

| Age/domain of assessment | NNU admissions (N=331) |  | NNU admissions >48h (N=218) |  | Preterm (N=142) |  |
| --- | --- | --- | --- | --- | --- | --- |
|  | OR (95% CI) | P value | OR (95% CI) | P value | OR (95% CI) | P value |
| Age 11 – Reading |  |  |  |  |  |  |
| Unadjusted | 1.23 (0.81-1.85) | 0.330 | 0.95 (0.59-1.53) | 0.843 | 1.58 (0.84-2.98) | 0.160 |
| Adjusted | 1.10 (0.68-1.77) | 0.695 | 0.76 (0.43-1.33) | 0.342 | 1.62 (0.83-3.19) | 0.160 |
| Age 11 – Writing |  |  |  |  |  |  |
| Unadjusted | 1.34 (0.91-1.96) | 0.127 | 1.29 (0.86-1.95) | 0.232 | 1.90 (1.06-3.41) | 0.030 |
| Adjusted | 1.18 (0.75-1.82) | 0.473 | 1.08 (0.67-1.76) | 0.741 | 1.92 (1.02-3.59) | 0.042 |
| Age 11 - Mathematics |  |  |  |  |  |  |
| Unadjusted | 1.54 (1.02-2.33) | 0.004 | 1.51 (1.01-2.25) | 0.043 | 2.10 (1.18-3.73) | 0.011 |
| Adjusted | 1.47 (0.96-2.25) | 0.061 | 1.24 (0.77-1.99) | 0.374 | 2.11 (1.13-3.93) | 0.018 |

Unadjusted odds ratios (ORs), adjusted odds ratios (aORs), 95% confidence intervals (CIs), p-values, and sample sizes are shown for each educational outcome. Outcomes represent not meeting the nationally expected standard at each assessment point. Models were adjusted for maternal, child, socio-economic, and school-level covariates. Children with no neonatal care exposure were the reference group.

The proportion of children not meeting expected educational standards was similar at ages 5 to 11. At age 5, 27% of children did not achieve expected educational attainment, and 20% at age 6 for the phonics assessment. At age 7, 24% did not meet expected standards in reading, 32% in writing, 24% in mathematics, and 18% in science. By age 11, 17% did not meet expected standards in reading, 21% in writing, and 25% in mathematics (S2A Table).

### The prevalence of special educational needs (SEN) at age 11 among NNU graduates

Within the NNU cohort, 12% of children were identified as requiring SEN support, with 3% holding an Education, Health and Care (EHC) plan. Among term-born children admitted to NNU for more than 48 hours, 11% were recorded as having SEN support, including 4% with an EHC plan. In the preterm cohort, 6% of children required SEN support or held an EHC plan.

Children who had experienced any NNU care were more likely to require additional educational support in mid-childhood (Table 2) (aOR, 1.78; 95% confidence interval [CI], 1.15-2.73; p=.009) compared to the study control group. This pattern remained when the analysis was restricted to term-born children who required longer NNU stay (>48 hours) aOR, 1.88; 95% CI, 1.19-3.00; p=.007), and to children born preterm (<37+0) without ongoing physical health problems (aOR, 1.50; 95% CI, 1.00-2.25; p=.049).

**Table 2:**
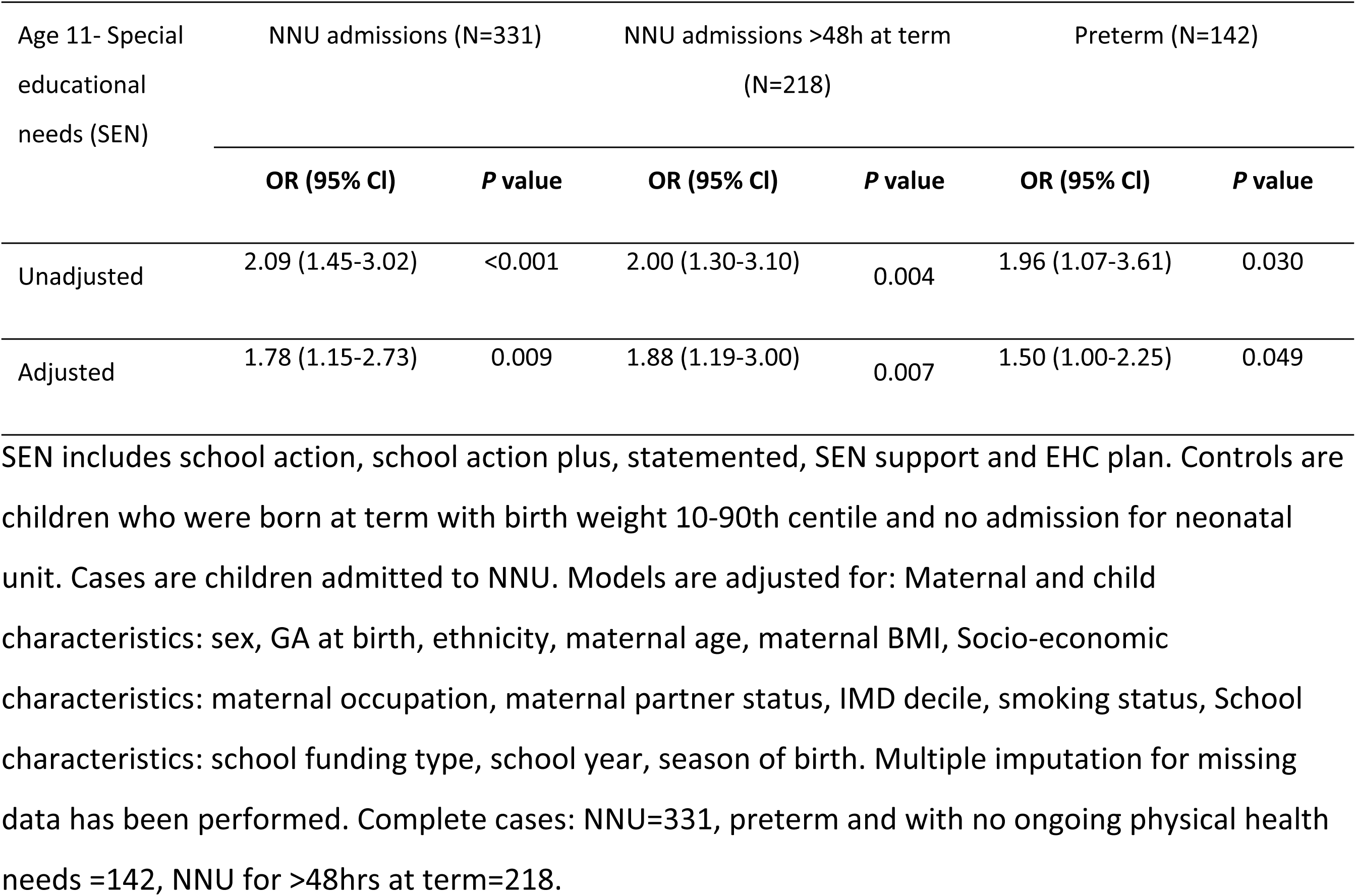
Special educational needs (SEN) among any NNU admission, > 48 hours NNU, or preterm babies without ongoing physical health needs.

| Age 11- Special educational needs (SEN) | NNU admissions (N=331) |  | NNU admissions >48h at term (N=218) |  | Preterm (N=142) |  |
| --- | --- | --- | --- | --- | --- | --- |
|  | OR (95% CI) | P value | OR (95% CI) | P value | OR (95% CI) | P value |
| Unadjusted | 2.09 (1.45-3.02) | <0.001 | 2.00 (1.30-3.10) | 0.004 | 1.96 (1.07-3.61) | 0.030 |
| Adjusted | 1.78 (1.15-2.73) | 0.009 | 1.88 (1.19-3.00) | 0.007 | 1.50 (1.00-2.25) | 0.049 |

SEN includes school action, school action plus, statemented, SEN support and EHC plan. Controls are children who were born at term with birth weight 10-90th centile and no admission for neonatal unit. Cases are children admitted to NNU. Models are adjusted for: Maternal and child characteristics: sex, GA at birth, ethnicity, maternal age, maternal BMI, Socio-economic characteristics: maternal occupation, maternal partner status, IMD decile, smoking status, School characteristics: school funding type, school year, season of birth. Multiple imputation for missing data has been performed. Complete cases: NNU=331, preterm and with no ongoing physical health needs =142, NNU for >48hrs at term=218.

SEN status was strongly associated with not meeting expected educational standards; however, the overlap between these measures was incomplete (S1 Fig). Three distinct groups were observed. A subset of children both required SEN support and did not meet expected standards, representing those with the greatest combined need. However, a comparable number of children with SEN achieved expected academic standards. Conversely, a separate group of children failed to meet expected standards in the absence of identified SEN.

Predictive performance of NNU admission for SEN at age 11 is reported in the sensitivity analyses in S4A-C Tables. Sensitivity analyses supported the distinction between educational attainment and educational support needs. A moderate discrimination for early educational outcomes across neonatal exposure groups was found, with AUC values ranging around 0.69-0.75 between ages 5 to 7 (S4A Table). In contrast, discrimination for age 11 academic attainment was weaker and less consistent (AUC 0.49-0.71), particularly for NNU admission alone, whereas discrimination for SEN remained moderate across all exposure groups: any NNU admission (AUC 0.70, S4A Table), NNU >48 hours at term (AUC 0.70, S4B Table), and preterm birth without ongoing physical sequelae (AUC 0.69, S4C Table).

## DISCUSSION

### Principal findings

NNU admission was consistently associated with an increased likelihood of poorer educational attainment in early mid-childhood and a sustained need for special educational support into mid-childhood. These associations were robust to adjustment for perinatal, demographic, and socio- economic factors and school funding model within the UK state system. By age 11, differences in academic attainment were attenuated and no longer clearly distinguishable from chance, whereas the association with SEN persisted. These findings suggest that the longer-term impact of NNU admission is more evident in the need for additional learning support than in overall academic performance. We demonstrate that while NNU admission, as a stand-alone metric has poor discriminative ability for predicting individual academic outcomes, there is potential to use this information within early risk-stratification pathways for SEN.

### Results in the context of what is known

These findings are consistent with previous literature demonstrating that early-life adversity, including prematurity and neonatal illness, is associated with later developmental and educational challenges [20–25]. Prior studies have shown that children requiring neonatal care are at increased risk of neurodevelopmental impairment and lower cognitive performance [26–29]. Our results extend this evidence by demonstrating that these risks are evident across multiple educational domains in early childhood, but that academic attainment may partially converge with peers over time. The persistence of increased SEN requirements aligns with emerging evidence that early-life health adversities may manifest as ongoing support needs rather than fixed deficits in attainment [30]. Alternatively, access to SEN support may itself help mitigate the impact of neonatal adversity on later educational attainment, such that support needs remain elevated even where attainment differences are less pronounced. The observation that associations persist even in children without ongoing physical health conditions suggests that these effects are not explained solely by prematurity or lasting medical complications, but may reflect broader developmental impacts of early-life illness. Furthermore, the incomplete overlap between SEN and academic attainment is consistent with prior work indicating that these measures capture related but distinct dimensions of child development. Together, these findings highlight the distinction between population-level associations and individual-level prediction, which has been less clearly addressed in previous studies. Of note, differences between the NNU and control groups were observable despite the cohort as a whole having lower SEN support than the general population. For context, national data for England report that 14.2% of pupils receive SEN support and 5.3% have an EHC plan [31].

### Clinical implications

NNU admission represents a simple early-life marker that is readily ascertainable by medical records and parental report and could be incorporated into routine assessments to identify children at increased risk of requiring additional educational support. Although it is not suitable as a standalone predictive tool for academic attainment, its consistent association with SEN suggests potential utility within broader risk stratification frameworks. Integration of NNU history into early years assessments by health visitors, primary care practitioners, and educational settings may support earlier identification of children who would benefit from enhanced monitoring or targeted interventions. The partial overlap between SEN and attainment further supports the need for multidimensional assessment approaches, rather than reliance on single outcome measures. This approach aligns with current priorities to improve early identification of developmental needs and reduce later educational inequalities.

### Strengths and limitations

Strengths of this study include the use of a large, well-characterised prospective birth cohort with detailed perinatal phenotyping and linkage to standardised, nationally collected educational outcomes. The use of routinely collected educational data enhances generalisability and relevance to real-world settings. The availability of detailed covariate information enabled robust adjustment for potential confounding factors, and findings were consistent across multiple domains and subgroups, supporting applicability across diverse educational contexts.

Several limitations should be considered. First, the cohort is derived from a single geographic region (Cambridge, United Kingdom) and consists of nulliparous women, largely of white ethnicity, which may limit generalisability to broader populations. Second, analyses at age 11 were based on a smaller subset of the cohort due to data availability, partly reflecting disruption to educational assessments during the COVID-19 period, resulting in reduced statistical power and wider confidence intervals. Third, although extensive covariate adjustment was performed, residual confounding cannot be excluded. Finally, while NNU admission captures a heterogeneous group of conditions, the analysis does not fully disentangle the relative contributions of underlying diagnoses or severity of illness. Nevertheless, the current model demonstrates that NNU admission represents a simple and readily ascertainable early-life marker associated with later educational attainment and SEN.

### Conclusions

NNU admission is associated with an increased risk of adverse educational outcomes in early childhood and a persistent requirement for additional educational support into mid-childhood, independent of perinatal and socio-demographic factors. These associations are observed even in the absence of ongoing physical health conditions. However, substantial heterogeneity in outcomes remains, and NNU admission alone lacks sufficient discriminatory capacity to predict individual developmental trajectories.

These findings emphasise the distinction between association and prediction. NNU admission should not be interpreted as a diagnosis of poor educational outcome, nor does it imply that children requiring neonatal unit are destined to experience adverse developmental trajectories. Rather, NNU admission represents a readily identifiable and pragmatic early-life marker of increased risk, with potential value within multivariable risk stratification frameworks.

In practice, incorporation of NNU history into routine early years assessments may support earlier identification of children who could benefit from enhanced monitoring or targeted intervention. Its accessibility and simplicity make it well suited for inclusion in low-burden screening approaches aimed at reducing subsequent educational inequalities.

## DATA AVAILABILITY

The POPS raw dataset is not publicly accessible due to the consent conditions under which participants originally agreed to take part. Requests for additional POPS materials, including raw data, may be directed to Mrs Sheree Green-Molloy at the Department of Obstetrics and Gynaecology, University of Cambridge, United Kingdom.

The educational information used in the study are derived from a customised extract of the National Pupil Database, containing anonymised individual-level results from routinely conducted national assessments. The National Pupil Database is jointly controlled by the UK Department for Education, Department for Work and Pensions, Higher Education Statistics Authority, and HM Revenue & Customs. For this study, access to the database was obtained through the Office for National

Statistics Secure Research Service under a data-sharing agreement with the University of Cambridge. Researchers can apply for access via the UK government application process: https://www.gov.uk/guidance/apply-for-department-for-education-dfe-personal-data.

## ACKNOWLEDGEMENTS

This research was funded by the NIHR Cambridge Biomedical Research Centre and supported by the NIHR Cambridge Clinical Research Facility (NIHR203312) and by the R&D Missions Accelerator Programme (R&DMAP) UKRI3106: Towards Neonatal Risk Stratification for Targeted Early Learning Support in the UK. The views expressed are those of the authors and not necessarily those of the NIHR or the Department of Health and Social Care.

## SUPPLEMENTARY INFORMATION (1)

## SUPPLEMENTARY INFORMATION FIGURE LEGENDS

**S2A Table: NNU, NNU >48 hours, preterm and mid-childhood academic outcomes.** Outcome: not achieving expected educational standard at each age/domain (as appropriate). ORs for unadjusted models and adjusted ORs for adjusted models with 95% CIs are displayed. Data show three cohorts: NNU (N=331), NNU >48 hours (N=269), and preterm babies without serious long-term medical sequelae (N=142); of these, N=77 also met criteria for exclusion of significant childhood morbidity in sensitivity analyses. Participants who had not experienced NNU, birth at term and healthy were the referent group . For adjusted models, covariates included in all models were the following: maternal age at pregnancy, maternal body mass index at recruitment, maternal ethnicity, maternal occupation, partner status, maternal smoking history, infant sex, gestational age at birth, birth weight standard deviation score, birth seasonality, childhood physical health, maternal index of multiple deprivation, school funding type, and academic year.

CI, confidence interval; NNU, neonatal unit; OR, odds ratio.

**S2B Table: Sex differences.** Models are adjusted for: Maternal and child characteristics: sex, gestational age at birth, ethnicity, maternal age, maternal BMI, Socio-economic characteristics: maternal occupation, maternal partner status, IMD decile, smoking status School characteristics: school funding type, school year, season of birth. Multiple imputation for missing data has been performed. Total for predictor of interest n=1281; Complete cases (female): EY=1189, Phonics=1181, Reading KS1=1024, Writing KS1=1024, Mathematics KS1=1024, Science KS1=1024, Reading KS2=589, Writing KS2=589, Mathematics KS2=589. Complete cases (male): EY=1193, Phonics=1197, Reading KS1=1052, Writing KS1=1055, Mathematic KS1s=1055, Science KS1=1055, Reading KS2=1135, Writing KS2=1135, Mathematics KS2=1135.

**S3 Table: Baseline characteristics of all eligible POPS participants and the analytic sample.** The full analytic sample is N=3423. Any low numbers below 10 are suppressed and made non-disclosive to comply with ONS data presentation policy. Data are obtained from the original dataset before imputation. For fields where there is no “missing” row, data were 100% complete. Maternal age was defined as age at recruitment. Maternal BMI was derived from weight measured at recruitment (12 weeks of gestation) divided by the square of height measured at 20 weeks of gestation (kg/m^2^). All other maternal characteristics were either self-reported at the 20 week-gestational age visit, from examination of the clinical record, or linkage to the hospital’s electronic databases. Deprivation was quantified using IMD 2007 based on census data from the area of the mother’s postcode. Birth weight percentiles and z scores were calculated using UK 1990 growth reference. SGA at birth is defined as birth weight <10^th^ percentile according to UK 1990 growth reference.

FTE indicates full-time education; IMD, index of multiple deprivation; MoM, multiple of the median; wk, week.

**S4A Table: AUC and sensitivity for risk stratification of NNU admission.** Area under the receiver operating characteristic curve (AUC), specificity, sensitivity, positive predictive value (PPV), and negative predictive value (NPV) for risk stratification of NNU admission across childhood educational outcomes and special educational needs (SEN). Performance metrics are shown for educational attainment outcomes assessed at ages 5, 6, 7, and 11 years. NNU admission status was evaluated against later educational outcomes using logistic regression-derived predicted probabilities. The referent group comprised children born at term, with birth weight between the 10th and 90th centiles, and without NNU admission. Analyses included children with available outcome data at each educational time point. AUC, area under the curve; NNU, neonatal unit; NPV, negative predictive value; PPV, positive predictive value; SEN, special educational needs.

**S4B Table: AUC and sensitivity for risk stratification of NNU >48 hours.** Predictive performance of prolonged NNU admission (>48 hours) for later childhood educational attainment and special educational needs (SEN), including area under the receiver operating characteristic curve (AUC), specificity, sensitivity, positive predictive value (PPV), and negative predictive value (NPV). Outcomes were assessed at ages 5, 6, 7, and 11 years using logistic regression-derived predicted probabilities. The referent group comprised children born at term, with birth weight between the 10th and 90th centiles, and without NNU admission. Analyses included children with available educational outcome data at each respective time point. AUC, area under the curve; NNU, neonatal unit; NPV, negative predictive value; PPV, positive predictive value; SEN, special educational needs.

**S4C Table: AUC and sensitivity for risk stratification of preterm births.** Predictive performance of preterm birth without significant childhood morbidity for later childhood educational attainment and special educational needs (SEN), including area under the receiver operating characteristic curve (AUC), specificity, sensitivity, positive predictive value (PPV), and negative predictive value (NPV).Outcomes were assessed at ages 5, 6, 7, and 11 years using logistic regression-derived predicted probabilities. Significant childhood morbidity referred to illnesses not considered directly related to intrauterine development that could independently influence educational attainment. The referent group comprised children born at term, with birth weight between the 10th and 90th centiles, and without NNU admission. Analyses included children with available educational outcome data at each respective time point. AUC, area under the curve; NNU, neonatal unit; NPV, negative predictive value; PPV, positive predictive value; SEN, special educational needs.

**S1 Fig:**
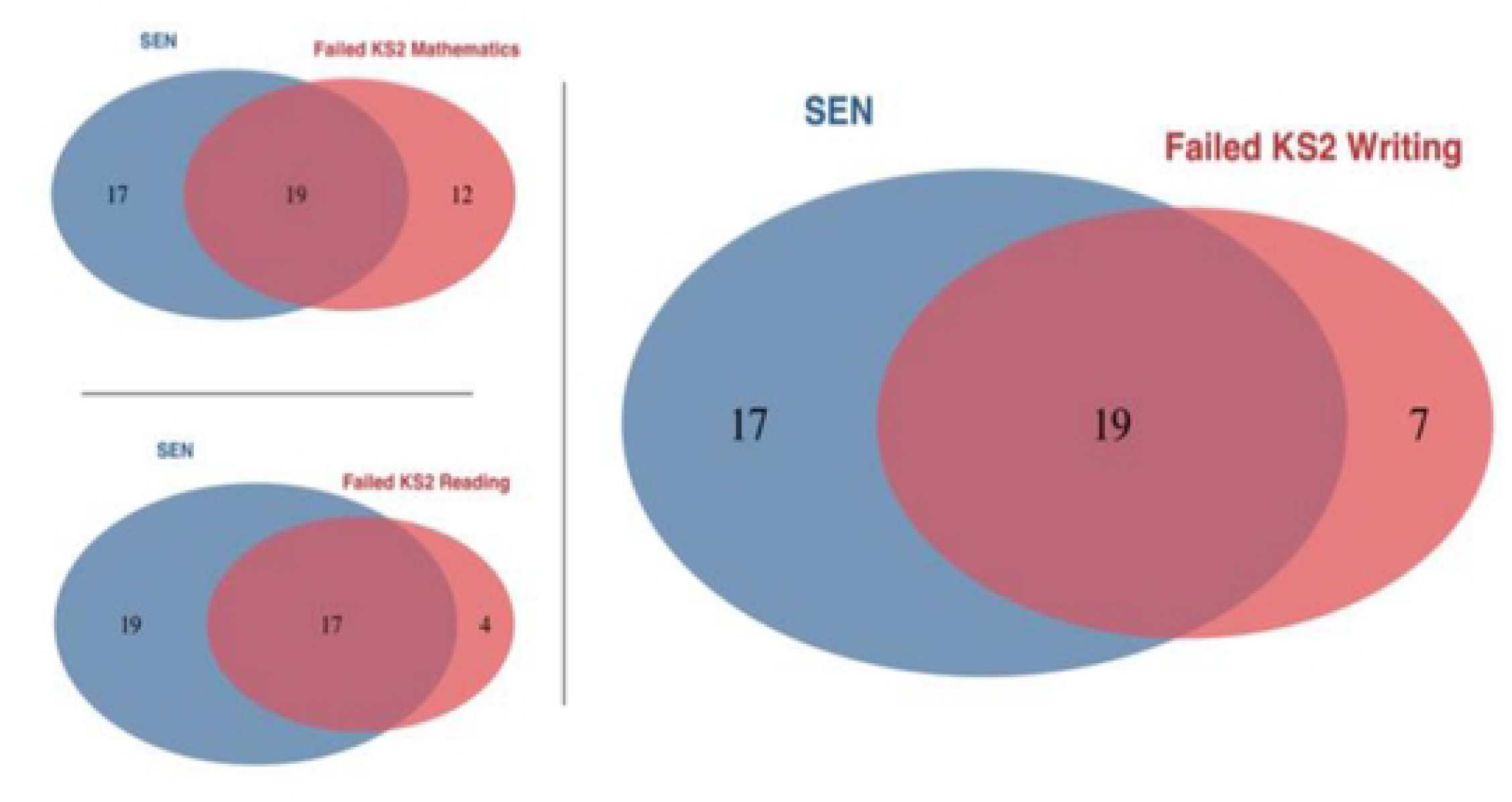
SEN and mid-childhood educational attainment. Venn diagram illustrating the overlap between SEN status and failure to meet expected standards in mathematics, reading, writing (GPS) within the NNU cohort. A substantial proportion of children who did not meet expected standards also had identified SEN, with a smaller number failing without SEN and some children with SEN achieving expected standards. This demonstrates that while SEN is strongly associated with poor reading attainment, it does not fully account for all cases of underperformance.

